# Trustworthiness of Systematic Review Automation An interview at Coventry University

**DOI:** 10.1101/2023.12.14.23299933

**Authors:** Xiaorui Jiang

## Abstract

To reduce both time and labour for systematic review, much effort has been made on developing automated tools using artificial intelligence and machine learning in nearly two decades. Unfortunately, systematic review automation tools still face a serious problem of social acceptance. Previous studies identified lack of trust as one important adoption barrier for systematic review automation. However, further discussion about building trust was limited to building trusted evidence base by benchmarked large-scale evaluation. This study extended the previous discussions of the trustworthiness of systematic review automation. Through semi-structure interviews with regular systematic reviewers, we tried to not only get answers for to what extent human reviewers trust automated tools and why, but also reveal more measures of building trust from human reviewers’ points of view and the impact of such measures on the trust in and adoption of systematic review automation tools. We believe that the results of this study may also shed light on some new directions of systematic review automation research.

## 1. Introduction

Systematic review (SR), sometimes called systematic literature review (SLR), is a prominent tool to establish a comprehensive understanding of a research topic. It is a necessary step towards scientific innovation that is widely adopted in many fields of study, such as medical and health sciences. Common SR steps include literature search, primary study selection (by title, abstract screening, full-text screening, and citation screening), quality assessment (of selected studies), data extraction (from selected studies), data synthesis and meta analysis, and summarisation and reporting (Tsafnat et al., 2014; also see Figure 1). Due to the huge number of candidate publications obtained by literature searching, the cost of doing a SR is extremely high in terms of both time and human labour (Michelson & Reuter, 2019).

**Figure 1.**
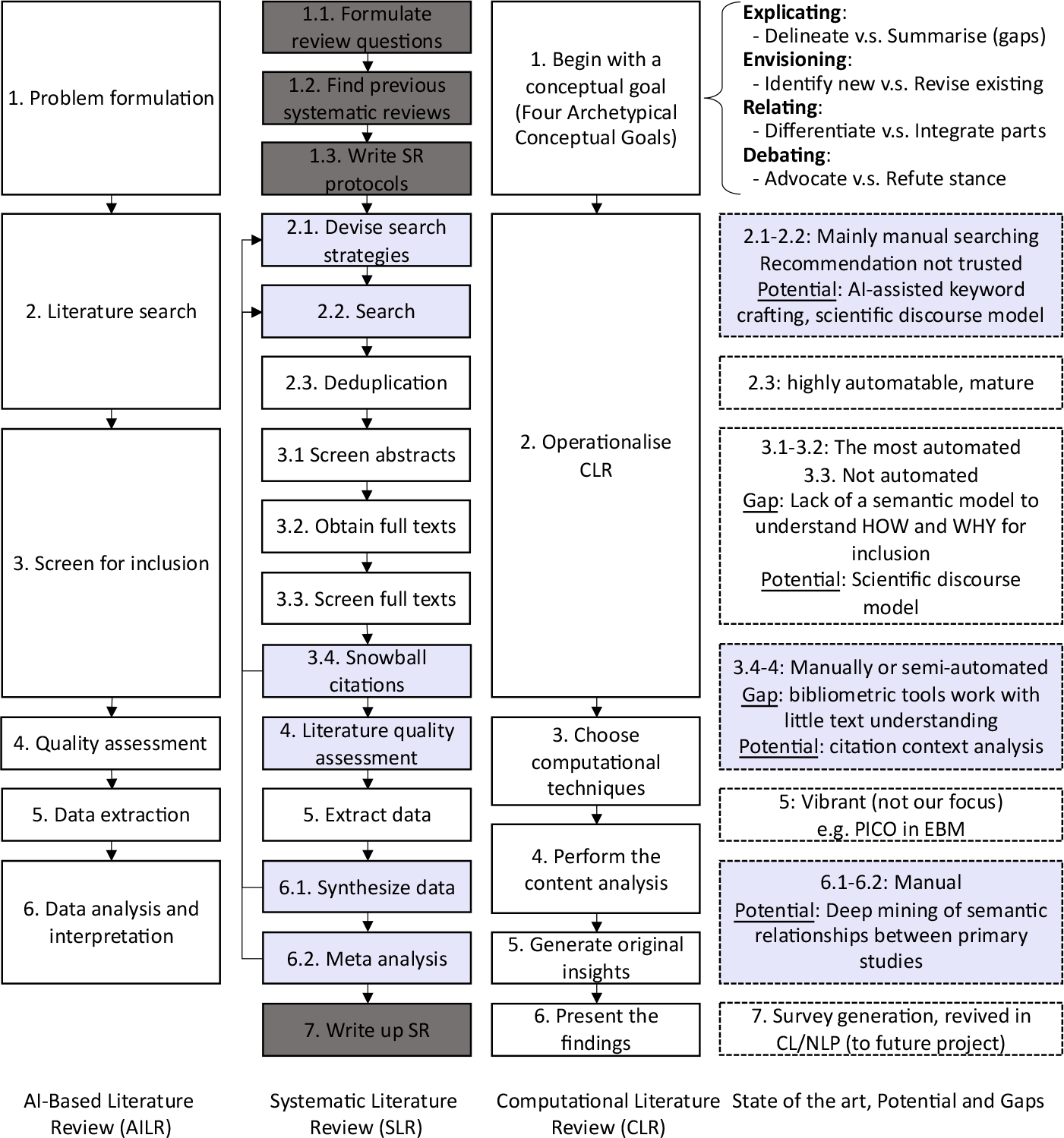
Flow chart of systematic review and gap analysis of two related AI-bases paradigms of review automation. Grey boxes represent the steps that are mainly done in a manual way.

A lot of efforts have been devoted to speeding up the SR process by automating or semi-automating the SR pipeline, mainly certain steps of the SR pipeline (Tsafnat et al., 2014). A wide range of automation tools have been developed in the past decade. While meta-analysis is the most successfully automated step, the majority of the research has been devoted to the selection of primary studies (van Dinter et al., 2021) because the number of publications to be scanned can be as large as tens of thousands. Meanwhile, while tools were mainly developed for medical and health sciences, recent years also witnessed similar paradigms arisen from other fields, such as Computational Literature Review (CLR; Antons et al., 2021) and AI-based Literature Review (AILR; Wagner et al., 2021). Figure 1 summarises and maps the core steps of the three literature review paradigms.

Despite efforts of nearly two decades, there are still adoption barriers for systematic review automation tools. Lack of trust was identified as the most significant factor hindering technology acceptance by the systematic review community (O’Connor et al., 2019b). The community of systematic review automation believed that his lack of trust rooted in the lack of evidence base that could demonstrate the “documented greater relative advantage”, i.e., “the degree to which an innovation is perceived as better than the idea it supersedes will affect its adoption” (O’Connor et al., 2018). The “importance of evaluation of automated tools” was recurringly identified as a major theme in the past meetings of International Collaboration of Automated Systemic Review (ICASR) (O’Connor et al., 2018, 2019a, 2020), which covers not only validity test on standardised benchmarks but also reproducible reporting of validity experiments (O’Connor et al., 2019b). Setup challenges and awareness of such tools were two other adoption barriers.

In this study, we aimed to justify the previously made claims about the trustworthiness issue of automated tools. To achieve this, we performed a series of semi-structured interviews with regular systematic reviewers who are from various research fields and not familiar with artificial intelligence technologies (i.e., who had no experience in developing artificial intelligence or machine learning tools by themselves). Taking a step further, this study also aimed to extend the previous study of the “quest for trust” (O’Connor et al., 2019b) by identifying new adoption barriers and investigating the routes leading to complete the quest for user trust. The findings of this study can be viewed as a companion to the gap analysis made by Hoang and Schneider (2022).

## 2. Methods

The research methodologies we applied were similar to Hoang and Schneider (2022). We conducted a series of semi-structured interviews with researchers in Coventry University who do systematic reviews (SR) regularly. After searching the PURE portal of Coventry University with “systematic review”, “systematic literature review”, “meta-analysis”, “qualitative review” in the titles of the publications, we identified more than 50 regular systematic reviewers with at least one published systematic reviewer papers, most with two or more. We sent out invitation emails to 50 researchers, including PhD candidates who were close to graduation at the time of invitations. We received about 15 positive responses and finally 10 were successfully invited to interviews within the required timeframe.

We used interviews to collect and investigate three categories of information: (1) the visibility of systematic review tools and systematic review automation (SR automation) to human reviewers (Q1-Q2 in Sect. 3); (2) the acceptance of SR automation to regular systematic reviewers, i.e. whether human reviewers trust automated tools, and the factors impacting such acceptance (Q3-Q4 in Sect. 3); and (3) what impacts the explainability of SR automation tools can have on their acceptance and what expectations end users have on explainable SR automation solutions.

The demography of participants shows a vert skewed distribution across fields, mainly because systematic reviewers mainly come from medicine, health, and life sciences domains (The “Field” column in Table 1). However, we tried our best to invite participants at a diverse range of career stages. The “Career stage” column in Table 1 shows the number of years of post-doctoral research experience in the academia, and “0” means PhD candidates close to graduation. Therefore, we have much universal distribution of career stage. This guaranteed a more comprehensive and diverse set of opinions about systematic review automation.

**Table 1.** Summary of Content Analysis of Interviews.

| No. | Career Stage | Subject | Experience with SR tools | Experience with SR automation | Expected steps to automate | % Time on searching & screening | Trust | Why trust/not trust<br>Factors impacting trust | After given explanations | Expectations from explanations (given to v.s. to give) |
| --- | --- | --- | --- | --- | --- | --- | --- | --- | --- | --- |
| P1 | 7+ | Bibliometrics | NVivo | No | Thematic analysis | n/a | NO | *Not conclusive, low recall and low precision*<br>n/a | Increased | Detailed report<br>n/a |
| P2 | 4-6 | Medicine | Ryyan, Covidence | No | Search, de-duplicate, screen | n/a | Hesitant | *Blackbox*<br>Piloted*; As safety check*; | Increased | Reasons/Rationales; Degree of certainty; PRISMA flowchart |
| P3 | 4-6 | Medicine | Refworks, NVivo | No | Search, de-duplicate, screen, quality assessment, meta analysis | 50% | Perhaps | *Accuracy, reproducibility*<br>Blended (human participation); endorsed (using it in publications); piloted | Better | Reasons based on eligibility criteria<br>n/a |
| P4 | 1-3 | Pharmacy | Endnote, Rayyan, Covidence | No | Search (across databases) | 50% | YES* | *Move forward*<br>Endorsed (using it in publications); safety check; Piloted | Innate | n/a<br>n/a |
| P5 | 7+ | Life sciences | Covidence, Rayyan | Yes? | n/a | 40-50%<br>(>30-40%) | Tend to | n/a<br>Piloted; Blended; Experiment/re-run | Better | Reasons<br>*Feedback (drop list based on criteria)* |
| P6 | 0 | Medicine | Covidence, NVivo | No | Screen, quality assessment, PRISMA, summary | 10%+40% | Tend to | *Machine dictates*<br>Safety-check; piloted; endorsed (by publications); Experiment/re-run | Better | Reasons<br>*Feedback (Yes/No)* |
| P7 | 3-6 | Health | NVivo | No | Search, screen (abstract plus full text), quality assessment | 50% | Perhaps | *Black box; "see it in action"*<br>Inspect decisions; Piloted; Experiment/re-run | Increased | Reasons; highlights; association between highlights and reasons<br>*Feedback based on criteria* |
| P8 | 0 | Medicine | Covidence, Rayyan, NVivo | No | n/a | 40% | Perhaps | *Black box; error*<br>Piloted (full recall, replicability) | Increased | Reasons aligned with criteria<br>*Feedback (checkbox based on criteria)* |

\* P8 responded with a strong YES but with conditions that the performance of SR automation tools were proved by pilot use

Our data analysis was partially rooted in thematic analysis. We manually corrected and aligned the auto transcripts generated by the meeting software, here Microsoft Teams, we used. Because we had predefined an initial set of questions to answer (detailed in Sect. 3.1), we first aligned the transcripts segments to these questions. Thematic analysis was applied to the non-aligned transcripts, allowing new themes to emerge. Then, we manually identified the codes used to perform content analysis of the interviews. In total, we had three themes as described above, each having two subthemes (detailed in Sect. 3.1), and one coding scheme to analyse each subtheme (detailed in Table 1.)

## 3. Interview Analysis

### 3.1. Focuses of Analysis

The following six questions were the focuses of analysis.

- Q1. The visibility of systematic review automation to academia.
- Q2. The visibility of systematic review automation tools to academia.
- Q3. The willingness of adopting/trusting systematic review automation tools.
- Q4. The factors that impact the trust in systematic review automation tools.
- Q5. The impact of explainability on the trust in systematic review automation tools
- Q6. End users’ expectations on the explainability of systematic review automation tools

Some research questions were predefined in the Interview Instructions (Appendix A). They are Q1, Q2, Q3, and Q5. For these questions, I manually picked out the longest segments of the interview transcripts and aligned the segments to one question. The coding scheme for the interview results of each question (here Q3 and Q5) was developed by clustering interview participants’ expressions that had a similar meaning.

The remaining two research questions, Q4 and Q6, were gradually emerged from thematic analysis of the remaining transcripts that followed the answers for Q3 and Q5 respectively. Corresponding transcript segments were manually aligned to each research question. The coding schemes for Q4 and Q6 were also developed by analysing the corresponding transcripts segments in a similar way as above.

### 3.2 Interview Results

Answers to each question that we intend to answer were coded in Table 1. Note that, two participants were finally found not regular systematic reviewers. They both came from the medical engineering subfield and had a stronger engineering background rather than medical and life sciences background. They shared a lot of valuable opinions about comprehensive literature review using in-house protocols and criteria that were not mappable to the common practice of systematic reviews. Therefore, we excluded their interview results because more than half of the columns in Table 1 could not be filled based on their interviews.

#### Q1. The visibility of systematic review automation to academia

The results are presented in the “Experience with SR automation” column. Unfortunately, although been developed for nearly two decades, *systematic review automation is almost totally invisible to human reviewers* in our context. Note that we are unable to conclude such invisibility to the whole academia because our data were only collected from Coventry University. We also want to emphasise that it was the concept of “systematic review automation” that was invisible to human reviewers due to the knowledge barrier between distinct fields like medicine and AI. However, some systematic review tools, even some with partial automatability, have already been used by many participants, without explicitly noticing such automation functionalities (detailed in the next subsection).

There was only one participant aware of automated systematic review tools. This was participant P5, the most senior researcher among all participants. Although P5 was less inclined to fully trust SR automation tools, this participant was one of the most positive and confirmative persons on the role of SR automation in systematic review in practice. An important opinion from P5 was that the more SRs a human reviewer does and the more frequently a human reviewer does SRs, the more inclined the human reviewer is to adopt SR automation in SR practice. P5 said:

> *P5: “So these people (i*.*e*., *PhD students), most of them … have experience in that level they … prefer do things manually. I feel like the colleagues that I’ve work, they they’ve done a systematic review here and there, …, They have a completely different view when you do like a proper systematic review like huge ones, and you do with a group of researchers. If I talk to any of my colleagues that they do this professionally, … there is no way that they will not use a software. … you cannot even think about [that]*.*”*

Witnessing such a huge knowledge gap, in the interviews, we had to spend roughly 10-15 minutes to introduce the concept of “systematic review automation” to all participants, explain how AI or machine learning works for certain SR steps, and advertise some existing SR automation tools that were used for nearly a decade or recently published. Interestingly, *all participants were quick at accepting the concept and overall responded positively to such tools*. This finding corroborates with conclusions of previous studies that lack of awareness and knowledge was a hinder of user trust in ST automation tools (O’Connor et al., 2019a,b).

#### Q2. The visibility of systematic review automation tools to academia

The answers for Q2 can be concluded from the “Experience with SR tools” column. In fact, *most participants had already used certain SR automation tools*, or at least certain SR tools that have some automation functionalities, *without awareness of the concept of SR automation*. For example, Rayyan (Ouzzani et al., 2016) is a tool that has the functionality to learn from human reviewers’ initial annotations to mark the relevant or irrelevant studies and highlight them in green/red colours^1^. Studies were also ranked using colour intensity. In other words, Rayyan has the functionality to automate the selection of primary studies, by automating the abstract screening step (Step 3.1 in Figure 1). Covidence^2^ is tool that can take in relevant and irrelevant studies annotated by human reviewers and automatically generate a PRISMA (Preferred Reporting Items for Systematic reviews and Meta-Analyses; Page et al., 2020; Rethlefsen et al., 2021) flowchart for reporting the systematic review process^3^. In fact, reporting the systematic review process is a mandatory requirement (mentioned by two participants) for publishing a systematic review, so the automatic generation of the PRISMA flowchart is an appealing feature.

We also noticed that *the range of tools that were used by the participants were quite limited*, almost confined to Rayyan, Covidence, and NVivo (for thematic analysis). Not a single automation tool for data extraction, data synthesis and meta-analysis was mentioned. This corroborates with the SR steps that our participants expected to automate with the help of AI/ML (see the “Expected steps to automate” column). These steps were mainly confined to literature search and citation screening (Step 2.1-3.3 in Figure 1). Participant P1 mentioned thematic analysis, which basically meant giving an understandable name to the word clusters generated by NVivo to help perform content analysis. From another side, most participants agreed that literature search and citation screening roughly take up 40-50% of the total time required to finish a systematic review (the “% Time on searching & screening” column). If we take into consideration the fact that usually at least two annotators are allocated for screening, a lot of time can be saved even if only citation screening is automated.

Two participants explicitly mentioned that data extraction is “fun” for human reviewers and that they were NOT bored by doing it manually. Two participants expressed their concern in the feasibility of automated data extraction and data synthesis. They thought these two are *high inference tasks* (O’Connor et al., 2020) and require very high-level cognitive capabilities that they did not believe current AI technologies could achieve. Finally, one participant (P3) pointed out the high potential of automated meta-analysis because this step only involves statistical analysis. The argument was right, but unfortunately no participant was aware of any such automation tools on the market. In fact, automated meta-analysis is the most mature tool in the systematic review automation tool stack. Similarly, no participant was aware of any SR automation tools from SR Step 4 to Step 7 in Figure 1, except the automatic generation of PRISMA flowchart by participant P6, which partially belongs to Step 7.

#### Q3. The willingness of adopting/trusting systematic review automation tools

From the “Trust” column, we were surprised to see that users were more willing to adopt, in other words more inclined to trust, SR automation tools than our initial expectation. Originally, the coding scheme included “NO”, “Partial”, and “YES”. To reflect the more subtle differences between different responses, it was extended to a 5-grade system: NO, Hesitant, Perhaps, Tend to, YES. “Hesitant” means “tend not to trust” even after deeper discussion about the ways to improve trust and the impact of explanations of machine learning decisions. “Perhaps” is weaker than “Tend to” because the former requires user interactions with SR automation tools, while “Tend to” means trust will be established if SR automations tools were piloted on certain human-written systematic reviews and/or their performances are proved good based on large-scale evaluations.

There was only one “NO” and one “Hesitant”. There were three “Perhaps” and two “Tend to”. More interestingly, there was one “YES” (P4). Participant P4 had an important comment on the acceptance of/trust in automation tools,

> *P4: “To be honest, I would trust it. … I think if we want to move forward in our systematic review, we have to have a solid automated system that do screening for us that we need to trust as a researcher. … And then if it’s been advised to use it as a researcher, I think I’m obligated to do it through the system because that will help*.*”*

Although P4 was so confirmative in answering the question about trust, a follow-up comment revealed some important and common ways of improving user trust – through **piloting** and **endorsement**, which will be detailed in the next subsection.

> *P4: “But I think to be able to trust it, I think we have to read a lot of publication around that the system and how accurate and sensitive it is*.*”*

Participant P6 left a similar comment although P6’s answer was classified as “Tend to”,

> *P6: “There are many things that’s been dictated by computers, and we don’t even question that. Why would we question this then?”*

#### Q4. The factors that impact trust in systematic review automation tools

The “*Why trust/not trust –* Factors impacting trust” column summarises the reasons why some participants did not trust automation tools (or equivalently did not feel comfortable with AI/ML substituting human decisions) and the ways to improve user trust in SR automation tools. The coding scheme includes 5 codes:

- Blended: Human participation is seen as a must in the SR process, e.g., SR automation tools return reasons for machine-made decisions, users give feedback to machine-made decisions. The man-machine interaction can be either one-way or two-way. If interaction is two-way, it is in nature reinforcement learning.
- Piloted: Good performance or close-to-human performance must be demonstrated through large-scale evaluation with obvious documented larger relative advantage in reducing human workload.
- Endorsed: There must be real-world use of SR automation tools by organisations or in publications (i.e., published systematic reviews using SR automation tools exist).
- Experimented/re-run: This is a very interesting feature suggested by three participants (P5, P6, and P7). It was a high-level interaction (human being blended with machine) where users may check machine-made decisions, make some changes, re-run the automation algorithm, and see what changes happen. “Experiment/re-run” is more than the trialability dimension of Innovation Diffusion Theory, which was used to analyse the acceptance of SR automation tools in Arno et al. (2021). The meaning of the latter is more about the availability of trial version.
- Safety-checked: On the contrary, safety-check was defined as a weaker way of being “Piloted” or “Blended” as only a sample of instances are required to be tested or checked by user.

An important finding, though likely to be biased towards our sample population, is that *it is possible to greatly increase user trust* and, *if certain requirements were met, most participants tend to give full trust in SR automation tools and adopt them in practice*. Most participants claimed that full trust would be established if SR automation tools were endorsed by other users in real-world SR practice. For some participants, the necessary requirement for them to improve trust was the ability to interact with SR automation tools, for example, by checking the reasons for machine-made decisions. In a sense, some participants would take SR automation tools as a partner to replace the role of the second annotator in citation screening or quality assessment. Finally, participant P6 raised a very interesting point. Different from most interviewees who wanted to teach SR automation tools to improve the performance by giving feedback, P6 also believed that SR automation tools could teach users in the early stage of doing an SR,

> *“If they provided the explanation to me and then I see this article to, maybe, I can even reflect on my eligibility criteria. So what is not well defined and what may be confusing. It would be a mutual learning process in the at the beginning*.*”*

For some participants, it was enough to establish high trust in SR automation tools if such tools had been piloted on a large number of human-written systematic reviews and proved good or close-to-human performance (e.g., ∼100% recall and high accuracy). Interestingly, all participants but P1 mentioned “Piloted” as an important way of increasing user trust. Indeed, large-scale evaluation has been a recurring important theme in the previous ICASR meetings (O’Connor et al., 2018, 2019a, 2020). Benchmarked evaluation ensures the reproducibility or replicability of existing studies, which was believed to be an important factor of user trust. Actually, participant P4 explicitly mentioned “reproducibility” as a factor for human reviewers to trust automated tools, although P4 was initially not so confident in adopting such tools. This corroborates with the importance of reproducibility that was reiterated in O’Conner et al. (2019b).

#### Q5. The impact of explainability on the trust in systematic review automation tools

The coding scheme for the impact of the measures for improving trust included three codes:

- Increased: Trust increased, but not necessarily to a level of full trust and willingness of adoption. For example, P1 still insisted that he needs a “detailed report” of all explanations (see the “Expectations from explanations” column) and needs to “go through all of them”. P2 was still “hesitant” to use such automation tools.
- Better: Trust will be increased to a level that the user is willing to adopt such tools in their own SRs and trust the result of SR automation tools if they are piloted or endorsed.
- Innate: This only applies to the few participants who have strong trust in SR automation tools based on AI/ML.

We could see that three participants (P2, P7, and P8) mentioned that machine learning being a “black box” was a reason for them to not trust SR automation tools. In the interviews some participants pointed out the importance of ability to interact with SR automation tools (P3 and P7). We further asked *what such interaction is purposed for*. In the direction from machine to human, the answer was **explanations** of the machine-made decisions. From the “After given explanations” column, we see all participants agreed that explanations would increase their trust in SR automation tools. We guess that, although not explicitly mentioned, other participants were also aware of the black box nature of automated tools, and this was the reason for *increased trust if the black box was opened by giving explanations*.

#### Q6. End users’ expectations on the explainability of systematic review automation tools

The results were summarised in the “Expectations from explanations” column. The coding scheme for the machine-to-human interaction included five codes:

- Degree of certainty: About the confident levels of machine-made decisions.
- Highlights: Highlighted text snippets, usually keywords or phrases, that are coloured to visualise their association to the inclusion and exclusion decisions.
- Reasons/Rationales: An advanced type of explanation detailing the reasons for decision-making, usually based on eligibility criteria, i.e., which inclusion criterion or exclusion criterion is matched.
- Reasons-Highlight Association: A more advanced type of explanation which provides evidenced reasons for machine-made decisions.
- PRISMA (flowchart): It is a mandatory requirement of publishing a systematic review in a peer-reviewed journal or a systematic review database. This means generating the explanation of the whole systematic review process, which is an advanced feature, in a graphical way, which may be enhanced by a summary paragraph.

The coding scheme for the human-to-machine interaction, i.e., user feedback, included the following codes:

- Feedback by YES/NO: Binary indicators of the correctness of machine-made decisions
- Feedback based on eligibility criteria: An advanced type of user feedback detailing why a machine-made decision is right or wrong. Two formats of feedback were mentioned: drop-down list (single selection) or check box (multiple selections).

First of all, all participants mentioned user interaction with SR automation tools in one way or another. The only exception was P4, but this was because we were unable to go into that depth in the interview due to running out of time. This corroborates one important finding from the 2017 ICASR meeting that “One general skepticism toward machine-assisted tasks automated reviews devoid of **valuable human control and input**” (O’Conner et al., 2019). Three participants (P5, P6, P7) mentioned an even more advanced way of interaction – experimenting with SR automation tools.

Overall, we can conclude that *systematic reviewers, the majority of whom are alien to computer science, mainly expect simple ways of feedback or interaction*. They did not expect giving complex forms of feedback to SR automation tools. Instead, they preferred simple feedback methods like a few button or mouse clicks. Because eligibility criteria are the rules human reviewers carefully craft and fully rely on in screening, the forms of feedback are better to be aligned with eligibility criteria. This is technically feasible because every well-written SR must define a rigid protocol and must register the protocol in a systematic review database before submission for review. The protocol must also be published together with the SR. Eligibility criteria are part of the protocol.

In the other way round, *human reviewers did not expect complex forms of explanations*: most expected highlights and reasons aligned with eligibility criteria. *Most did not expect the complex reasoning process behind a decision*, although some participants mentioned that a decision is usually made by checking the criteria one by one and thus the decision process works like a decision tree in machine learning. However, we noticed that it was not because human reviewers did not want complex explanations, such as a full sentence explaining the reasoning process. Instead, it was because *human reviewers were unable to imagine that existing AI technologies are capable of generating complex forms of explanation in human language*, due to their lack of knowledge in AI. For example, one participant (P7) commented below.

> *“I can’t imagine how you’d. I imagine that’s a huge undertaking to then take those decisions and then get a natural, you know, a sentence that people would understand for every single thing, for every single project*.*”*

## 4. Concluding Remarks

In this study, we summarised the results of eight interviews with regular systematic reviewers from Coventry University. We focused on getting a better understanding of several important questions around the adoption of systematic review automation tools. These questions include to what extent human reviewers without a computer science background are familiar with systematic review automation (Q2) and automated tools (Q1), to what extent human reviewers are willing to trust and adopt automated tools in systematic review practice and why (Q3), and what measures may be able to improve the trustworthiness of systematic review automation (Q4). As a further step, we rephrased the machine-to-human feedback as explanations of machine-made decisions and tried to answer questions around the impact of explainability on trust in systematic review automation (Q5) and the measures towards better explainability from the perspectives of human reviewers (Q5).

Concerning the first two questions, most human reviewers were not familiar with the concept of systematic review automation/automated systematic review, even though most participants used at least one systematic review tool that has a significant automation functionality, e.g., Rayyan for citation screening or Covidence for PRISMA flowchart generation. This reveals the huge knowledge gap between developers and users of automated tools. Thus, it was not only the difficulty of set-up (O’Conner et al., 2019b), but also the lack of knowledge in AI-based automated tools (Scott et al., 2021) and the lack of visibility of automated tools to the community (van Altena et al., 2019) that hindered the adoption of systematic review automation. The toolbox used by our participants were extremely limited. On the one hand, we believe this was a severe data bias (e.g., different from the statistics obtained by Scott et al. (2021)) because our participants all came from the same university, many came from the same research centre, and the university only bought licences for a limited number of tools. On the other hand, we believe this fact somehow reflected the reality of low visibility of most automated tools that were developed in the past decade (van Altena et al., 2019).

Despite the lack of knowledge and awareness of systematic review automation, we saw a much higher potential to trust and adopt automated tools (Q3). However, this trust was not without cost. Participants pointed out five ways of building trust (Q4), of which benchmarked large-scale evaluation (coded as “Piloted”) was the No. 1 method mentioned by all. This confirms to the reiterated importance of evaluation in previous meetings of International Collaborations on Automated Systematic Reviews (O’Conner et al., 2018, 2019a, 2020). This study identified four more ways of improving trust: “Endorsed” by organizational use and by use in published systematic reviews, “Safety-checked” as a weakened version of “Piloted” on a sample of candidate studies; “Experimented” by adjusting machine learning algorithm’s behaviour, and “Blended” which means human-machine interaction.

More in-depth interview questions about the “Blended” way revealed that its real meaning was that machines give and present explanations to human reviewers in a certain form and allow users to give feedback on machine-made decisions and machine-generated explanations. All participants agreed that the explainability of automated tools would increase user trust, though not necessarily to a level of fully trusted adoption of such tools (Q5). Participants’ expectations for the form of interaction/explanation/feedback were “low”, compared to our original anticipation. Users expected simple ways of interaction, such as matched eligibility criteria as explanation and/or highlighted text snippets as evidence, but not comprehensive explanations in natural language sentences or the complex reasoning process behind the decisions (Q6). These findings show that explainable systematic review automation is a new direction worth of and in need of studying. Meanwhile we have to announce that such “low” expectations might be due to human reviewers’ lack of knowledge of modern AI’s capability, which limited their imagination. Anyway, the findings about Q6 were not entirely conclusive, on which we foresee future interviews in more depth.

## Data Availability

All data produced in the present work are contained in the manuscript

## Appendices

### A. Interview Instructions

Total time: 60 minutes to 90 minutes.

Part 1. Show the **diagram** about the alignments between SLR (Systematic Literature Review), AILR (Artificial Intelligence based Literature Review), and CLR (Computational Literature Review), ask the interviewee whether this matches his/her own experience in doing systematic review. Ask him/her to describe the **way** he/she does systematic review and the **difficulties** he/she met.

Ask the participants to share their estimation of how much time they spent on each stage, and their need for automating or semi-automating some steps of them using AI/ML.

Part 2. Ask the interviewee to talk about he/she did **literature search**, and what he/she thinks about the potential of improvement and anticipate the way AI/ML can help. Similarly, ask the interviewee to talk about he/she did the **selection of primary studies**, i.e., **screening**, and how he/she **assess the quality** of study.

Part 3. Based on his/her response, stimulate the interviewee to discuss about the **trust issue** of AI/ML tools and their generated results, e.g., how they trust the AI-suggested search keywords, how they trust the returned papers and the rankings of the papers, etc.

Part 4. Based on his/her response, let the interviewee further discuss what is the most comprehensive way of **presenting explanations** to the AI/ML results. Is it in short natural language explanations? Should a group of results be given the same/quasi-same explanation based on semantic clustering?

Part 5. Ask the interviewee to talk about his ideas about some other aspects of SLR automation and explanation, such as the aspects listed below, if they have not been touched and time permits:

- explaining the **relationships** between selected primary studies,
- explaining the domain **evolution** among selected primary studies,
- explaining the **reproducibility** of SLR automation, i.e., by automatically generating PRISMA statement diagram and the textual descriptions of the PRISMA statement.
- Explaining how well the SR automation tools perform by **large-scale evaluation**, for the purpose of persuading more people into using such tools
- Etc.

## Footnotes

1 https://www.rayyan.ai/

2 https://www.covidence.org/

3 There are other more proposals of systematic review protocols, such as the PRIOR (Preferred Reporting Items for Overviews of Reviews) statement (Gates et al., 2022)

